# Accessing AI mammography reports impacts patient interest in pursuing a medical malpractice claim: The unintended consequences of including AI in patient portals

**DOI:** 10.1101/2024.12.27.24319688

**Authors:** Elizabeth C. Song, Michael H. Bernstein, Parker S. Lay, Leo Druart, Elizabeth H. Dibble, Ana P. Lourenco, Grayson L. Baird

## Abstract

**Background:** Artificial intelligence (AI) tools are increasingly used in breast imaging and radiology more broadly. Patients express varying levels of trust and acceptance toward the incorporation of AI tools, although no research has examined how it can best be communicated in the patient portal setting.

**Methods:** English-speaking US women with 1+ prior mammogram were recruited via Prolific and randomized to one of thirteen conditions. All participants were shown a vignette asking them to imagine receiving a BI-RADS 1 (Negative) radiologist report from their patient portal. Participants in twelve conditions also received an AI report with one of four AI abnormality scores (not flagged: 0, 29; flagged: 31, 50) and 0-2 accompanying features (nothing; a only; a & b): (a) an abnormality cutoff threshold; (b) the AI tool’s False Discovery Rate (FDR) or False Omission Rate (FOR). As the primary outcome, participants indicated whether they would consider a lawsuit if a one-year follow-up found evidence of Stage 3 breast cancer. Secondary outcomes included hypothetical decisions regarding follow-up (e.g., second opinions), concern for breast cancer, and desire for additional imaging.

**Results:** Participants (n=1,623) were more likely to consider a lawsuit when AI was (versus was not) provided, p=0.001. However, for most AI abnormality scores, providing the abnormality cutoff threshold and FDR/FOR reduced lawsuit consideration relative to the AI abnormality score alone. Concern for breast cancer, desire for additional imaging, and follow-up requests (same radiologist, different radiologist, and ordering physician) increased as the AI abnormality score increased, though this was also often mitigated by providing the FDR (and sometimes FOR).

**Conclusion:** Disclosing AI feedback used for medical decision-making will impact patients’ perceptions and behaviors pertaining to malpractice and follow-up. Best practices are needed to engage and inform patients about the application of AI tools in their care while minimizing its unintended negative consequences.

## INTRODUCTION

Artificial intelligence (AI) tools in breast imaging have rapidly increased, offering significant potential for improving breast cancer detection and management. The Food and Drug Administration (FDA) has approved several AI tools for screening mammography with applications in triaging exams, detecting and classifying lesions, and assessing breast density.^1^ With increasing imaging volume and continued demand for breast cancer screening, radiomics and AI studies have demonstrated that AI-enhanced workflows hold promise for improving diagnostic accuracy and efficiency.^2^

Nonetheless, successfully implementing AI tools in clinical practice requires participation from all stakeholders, including radiologists, developers, policymakers, and patients.^3^ Given that screening examinations can cause anxiety, understanding patients’ reactions to the incorporation of AI in this space is crucial. Patient surveys reveal various levels of trust and acceptance toward AI tools in breast imaging.^4^ While many express positive attitudes toward radiologist and AI collaboration, concerns surrounding trust, transparency, and accountability persist.^4–6^ Critically, little research has examined patients’ perceptions of AI use for breast imaging—or imaging, more broadly—with respect to how it is communicated in a patient portal setting.

With the movement for transparency and patient-centered care, patient portals have played an increasing role in healthcare delivery, enabling patients to send messages, schedule appointments, and access lab and imaging results.^7^ As more AI tools are implemented, it is important to establish best practices for communicating these results to patients. For a traditional radiologist report, writing with patients as the audience—such as by reducing medical jargon—can facilitate communication and comprehension.^8^ For an AI-interpreted imaging report, however, there are additional considerations about effectively presenting information and ensuring patient autonomy.^9^ Put differently, while disclosing AI use in and of itself is important from an ethical standpoint, insufficient context provided to patients for how to interpret AI information could have unintended consequences.

The goal of this study was to investigate what contextual information should be provided to patients regarding a hypothetical screening mammogram. We compare participants’ hypothetical decision-making and attitudes when presented with a Breast Imaging Reporting and Data System (BI-RADS) 1 radiologist report (a report concluding no breast cancer was found) and variations of an AI-interpreted report. Our specific hypotheses are listed in Supplemental Table 1.

## MATERIALS & METHODS

### Participants and Setting

Participants were recruited online through Prolific and were eligible if they identified as a female, English-speaking adult living in the United States (US) with 1+ prior mammogram. In total, n=1,623 participants were included (n=361 and 802 failed a comprehension and manipulation check, accordingly). Demographic information is shown in Table 1.

**Table 1.** Demographic characteristics.

| **Characteristic** | **Median** | **IQR** |
| --- | --- | --- |
| Age | 51 | 46-59 |
| Number of Lifetime Mammograms | 5 | 2-10 |
|  | **n** | **%** |
| Race |  |  |
| American Indian or Alaska Native | 9 | 0.56 |
| Asian | 54 | 3.33 |
| Black or African American | 237 | 14.63 |
| More than one race | 49 | 3.02 |
| Native Hawaiian or Other Pacific | 1 | 0.06 |
| White | 1259 | 77.72 |
| Prefer not to say | 11 | 0.68 |
| Ethnicity |  |  |
| Hispanic or Latino/a | 96 | 5.94 |
| Not Hispanic or Latino/a | 1502 | 92.95 |
| Prefer not to say | 18 | 1.11 |
| Family Diagnosed Breast Cancer |  |  |
| Yes | 396 | 24.41 |
| No | 1152 | 71.02 |
| Unsure | 64 | 3.95 |
| Prefer not to say | 9 | 0.55 |
| Diagnosed Breast Cancer |  |  |
| Yes | 58 | 3.57 |
| No | 1557 | 95.93 |
| Prefer not to say | 8 | 0.49 |

### Procedure

Participants who screened and provided online consent were directed to an online Qualtrics survey where they were randomized to one of 13 conditions. Participants were compensated $1.25 for completing the survey and $0.14 if they screened out for never having a mammogram. Participants were prevented from participating more than once. Ethics committee/IRB of Brown University Health gave ethical approval for this work (IRB009824).

### Vignette Design and Experimental Conditions

All participants were shown a vignette asking them to imagine they had recently received their annual screening mammogram and logged into their patient portal to access the results. In all conditions, the hypothetical results consisted of a standard radiologist’s report with a BI-RADS 1 (Negative) determination and a 2D mammogram. No additional information was provided in the control condition.

In the remaining twelve conditions, participants were provided an additional AI-interpreted report with one of four abnormality scores: 0 (Not flagged for abnormality), 29 (Not flagged for abnormality), 31 (Flagged for abnormality), or 50 (Flagged for abnormality). Moreover, there were three conditions for each abnormality score: (1) only the abnormality score was presented, (2) both (1) and the abnormality cutoff threshold of 30 with a range 0-100 were presented, or (3) both (1) and (2) were presented in addition to either a) False Discovery Rate (FDR, complement of the positive predictive value, 1-PPV) for the abnormality score=31 or 50 conditions or b) False Omission Rate (FOR, complement of the negative predictive value, 1-NPV) for the abnormality score=0 or 29 conditions (Supplemental Table 2). For all 12 AI conditions, the 2D mammogram included the corresponding AI information (see Appendix II for full vignettes).

To maximize ecological validity, the hypothetical AI’s performance was based on a commercially available AI mammography interpretation software, Lunit INSIGHT MMG version 1.1.7.1 (Lunit, Seoul, South Korea).^10^ Using the optimal threshold of 30 with 72.38% sensitivity and 92.86% specificity described by Seker et al.—and assuming a breast cancer prevalence of 0.57%—the corresponding FDR and FOR were 95% and 0.2%, respectively. These were the FDR and FOR values given and explained to participants.^10^

### Measures

#### Checking behavior (i.e., behavior to reduce anxiety)

Four questions probed hypothetical decisions participants would make after receiving the mammogram report (yes/no): Would you request 1) “a second opinion from another radiologist?” 2) “a follow-up meeting with the radiologist who wrote your original report?” 3) “a follow-up consult with your primary care physician (including OB/GYN)?” and 4) “follow-up imaging that is more accurate even if a physician says it is not necessary for you?”

#### Litigation

As the primary outcome, to measure participants’ inclination to sue the radiologist, participants were told, “Imagine one year later you receive a new mammogram with a new report from a different radiologist that indicates evidence of potential breast cancer. A biopsy confirms that breast cancer is present and is stage 3 (which indicates advanced cancer). Would you consult an attorney to explore suing the original radiologist for not having found the cancer in the previous year?” (yes/no).

#### Trust

Two questions measured participants’ trust in the decisions of the radiologist and AI system depicted in the vignette, from 0 (not at all) to 100 (very much): “Do you ultimately trust…” 1) “the radiologist’s decision?” 2) “the AI’s decision?”

#### Concern

One question measured concern for breast cancer: “How, if at all, would these results change your concern about having breast cancer?” (−50-Greatly Decrease; 0-No Change; 50-Greatly Increase).

#### Comprehension and Manipulation Check

All participants had to correctly identify “No mammographic evidence of malignancy” as a comprehension check. All in AI conditions had to correctly recall the abnormality score (0, 29, 31, 50) and “(Not)flagged for abnormality” determination. Those in the cutoff conditions had to recall the correct cutoff of “30.” Those provided a FOR or FDR had to correctly recall “The AI had a False Omission Rate of 0.2%” or “The AI had a False Discovery Rate of 95%”, respectively.

### Statistical Analyses

All analyses were conducted using SAS Software 9.4 (SAS Inc. Cary, NC). Attitudes by experimental condition were modeled using generalized linear modeling assuming a normal or binary distribution, where appropriate with the GLIMMX procedure using contrast comparisons reflecting the hypotheses. All interval estimates were calculated for 95% confidence and alpha was established at the .05 level. P-values were adjusted for FDR correction using the Benjamini-Hochberg method.

## RESULTS

### Primary outcome

#### Litigation

As seen in Figure 1 and Supplemental Tables 3 and 4, consideration of a lawsuit after a BI-RADS 1 determination upon discovering breast cancer the following year increased when AI results were included relative to No AI, p=0.001. Consideration of a lawsuit increased from 39.7% with No AI (control) to 46.9% when the abnormality score of 0 was presented alone. However, this decreased back to the control of 40.0% upon providing both the cutoff threshold of 30 and the FOR of 0.2%—all other conditions where AI agreed with the radiologist had higher percentages pursuing legal liability compared to control, p=0.37. Once AI flagged the case as abnormal, consideration of a lawsuit increased to 60.7% when an abnormality score of 31 was provided alone, decreasing to 56.1% when paired with the cutoff threshold of 30. This decreased even further to 31.6% when the FDR of 95% was provided, p=0.0003. Moreover, when the abnormality score of 50 was presented alone, consideration of a lawsuit was 48.3%, which increased to 51.9% when paired with the cutoff threshold, which decreased to 36.8% when the FDR was provided, p=0.02.

**Figure 1.**
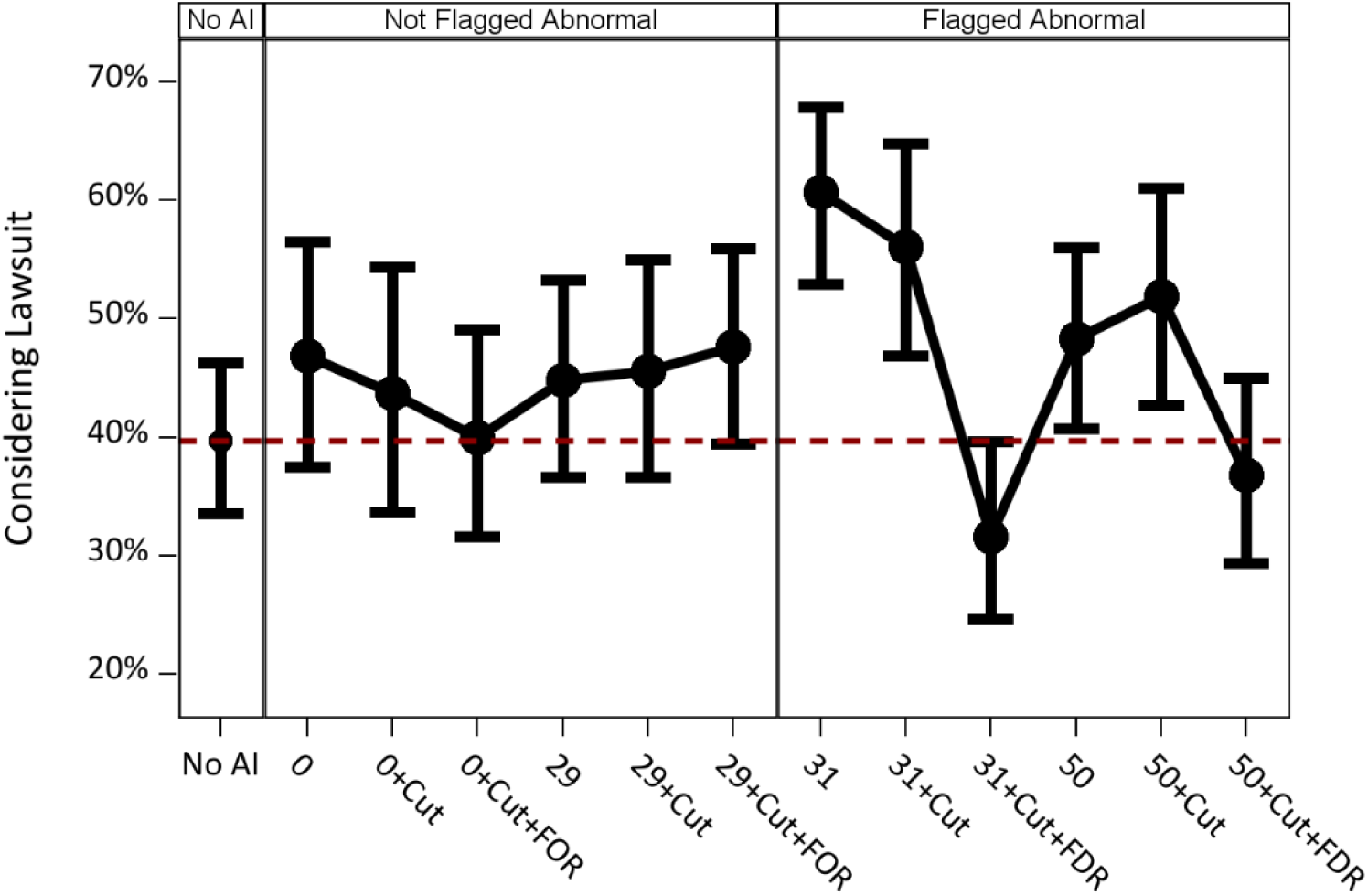
Considering a Lawsuit after a BI-RADS 1 determination upon discovering breast cancer the following year. X-Axis is experimental condition, broken down by three column panels (No AI, Not Flagged for Abnormality, Flagged for Abnormality). The 12 AI conditions are comprised of four abnormality scores (0, 29, 31, 50) crossed with: 1) the abnormality cutoff threshold of “30” (Cut), and 2) the error rates (FOR, false omission rate, .2%; FDR, false discovery rate, 95%). Y-axis is percentage of participants who indicated they would consult an attorney to consider suing the radiologist for the BI-RADS 1 interpretation. Red dashed line is control reference. Black dots are means with 95% confidence intervals.

### Secondary outcomes

#### Second opinion

As seen in Figure 2A and Supplemental Tables 3 and 5, desire to follow up with a different radiologist (second opinion) after a BI-RADS 1 determination increased when AI results were included relative to No AI, p=0.01. Desire for a second opinion increased from 8.3% with No AI (control) to 27.2% when the abnormality score of 29 was presented alone, which increased to 68.0% when paired with the cutoff threshold of 30, which decreased slightly to 62.5% when the FOR of 0.2% was included— note, the latter two were relatively high even though AI agreed with the radiologist. Once AI flagged the case as abnormal, desire for a second opinion increased to 70.2% when an abnormality score of 31 was provided alone and increased to 81.3% when paired with the cutoff threshold of 30, p=0.0003. This decreased to 61% when the FDR of 95% was provided, p=0.009. Likewise, when the abnormality score of 50 was presented alone, desire for a second opinion was 68.9%, which increased to 74.0% when paired with the cutoff threshold, which decreased to 60.9% when the FDR was provided, p=0.02.

**Figure 2 A-B.**
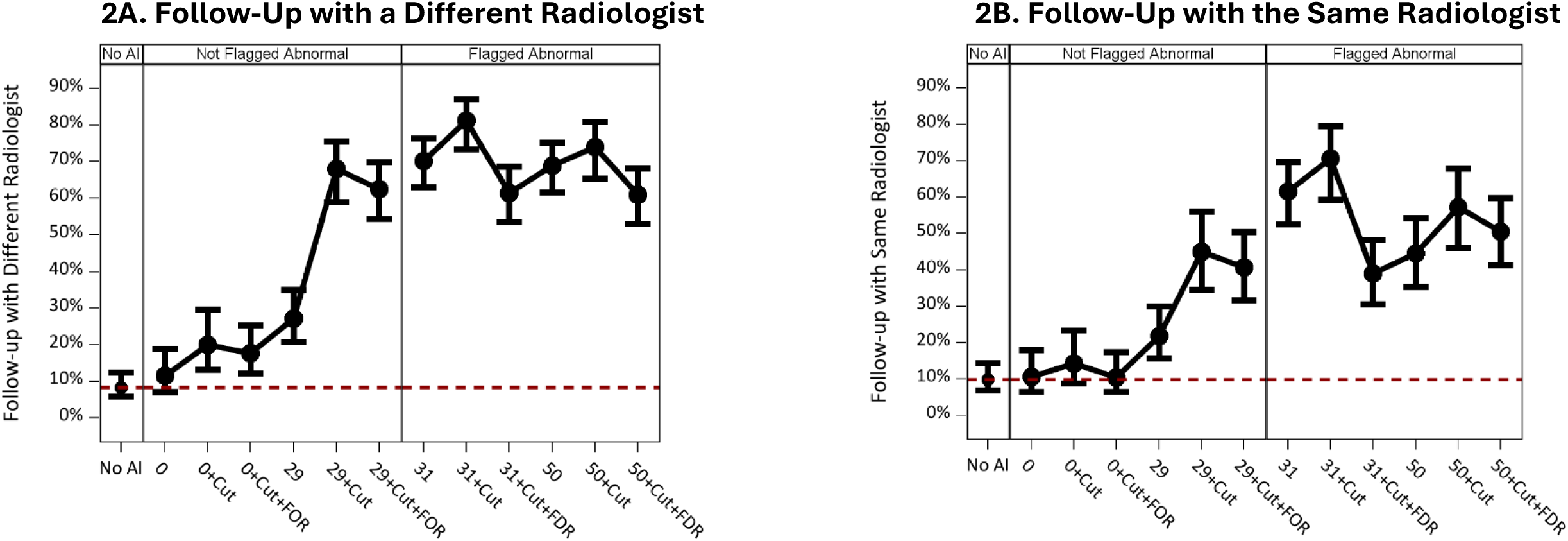
Follow-up Seeking behaviors. X-Axis is experimental condition, broken down by three column panels (No AI, Not Flagged for Abnormality, Flagged for Abnormality). The 12 AI conditions are comprised of four abnormality scores (0, 29, 31, 50) crossed with: 1) the abnormality cutoff threshold of “30” (Cut), and 2) the error rates (FOR, false omission rate, .2%; FDR, false discovery rate, 95%). Y-axis is percentage of participants who indicated they would pursue the follow-up behavior (0-100%). Red dashed line is control reference. Black dots are means with 95% confidence intervals.

#### Second look

As seen in Figure 2B and Supplemental Tables 3 and 5, desire to follow up with the same radiologist (second look) after a BI-RADS 1 determination increased when AI results were included relative to No AI, p=0.0002. Desire for a second look increased from 9.8% with No AI (control) to 21.8% when the abnormality score of 29 was presented alone, which increased to 45.1% when paired with the cutoff threshold of 30, which decreased slightly to 40.7% when the FOR of 0.2% was included—note, the latter two were relatively high even though AI agreed with the radiologist. Once AI flagged the case as abnormal, desire for a second look increased to 61.5% when an abnormality score of 31 was provided alone, increasing to 70.6% when paired with the cutoff threshold of 30, p=0.0002. This decreased to 39.0% when the FDR of 95% was provided, p=0.0002. Likewise, when the abnormality score of 50 was presented alone, desire for a second opinion was 44.6%, which increased to 57.4% when paired with the cutoff threshold, which decreased to 50.5% when the FDR was provided, but failed to reach statistical significance, p=0.21.

#### Primary care physician (PCP) follow-up

As seen in Figure 3A and Supplemental Tables 3 and 5, desire to follow up with the PCP after a BI-RADS 1 determination increased when AI results were included relative to No AI, p=0.02. Desire for PCP follow-up increased from 19.8% with No AI (control) to 37.6% when the abnormality score of 29 was presented alone, which increased to 67.7% when paired with the cutoff threshold of 30, which decreased slightly to 58.0% when the FOR of 0.2% was included— note, all three were relatively high even though AI agreed with the radiologist. Once AI flagged the case as abnormal, desire for PCP follow-up increased to 64.9% when an abnormality score of 31 was provided alone, increasing to 73.3% when paired with the cutoff threshold of 30, p=0.0002. This decreased to 55.3% when the FDR of 95% was provided, p=0.02. Likewise, when the abnormality score of 50 was presented alone, desire for PCP follow-up was 59.2%, which increased to 63.0% when paired with the cutoff threshold, which decreased to 58.8% when the FDR was provided, but failed to reach statistical significance, p=0.33.

**Figure 3 A-B.**
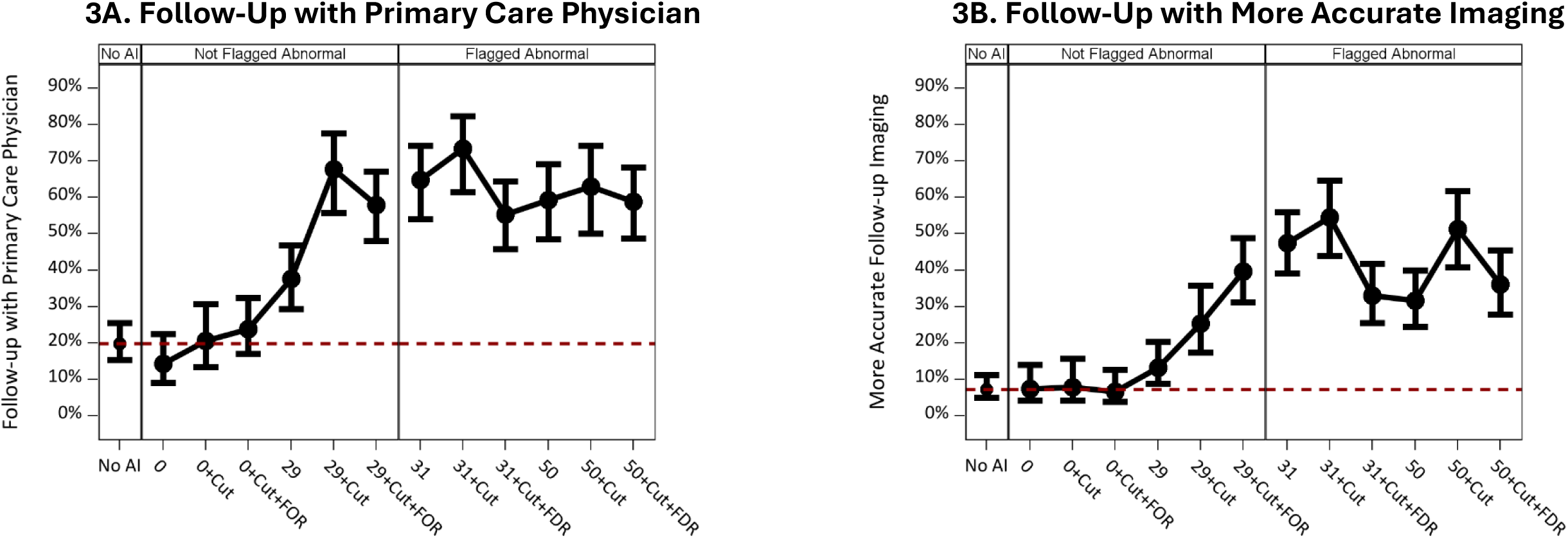
Follow-up Seeking behaviors. X-Axis is experimental condition, broken down by three column panels (No AI, Not Flagged for Abnormality, Flagged for Abnormality). The 12 AI conditions are comprised of four abnormality scores (0, 29, 31, 50) crossed with: 1) the abnormality cutoff threshold of “30” (Cut), and 2) the error rates (FOR, false omission rate, .2%; FDR, false discovery rate, 95%). Y-axis is percentage of participants who indicated they would pursue the follow-up behavior (0-100%). Red dashed line is control reference. Black dots are means with 95% confidence intervals.

#### Request for more accurate follow-up imaging (MAFI)

As seen in Figure 3B and Supplemental Tables 3 and 5, desire for MAFI after a BI-RADS 1 determination increased when AI results were included relative to No AI, p=0.0002. Desire for MAFI increased from 7.2% with No AI (control) to 13.3% when the abnormality score of 29 was presented alone, which increased to 25.3% when paired with the cutoff threshold of 30, which increased to 39.6% when the FOR of 0.2% was included—note, the latter two were relatively high even though AI agreed with the radiologist. Once AI flagged the case as abnormal, desire for MAFI increased to 47.5% when an abnormality score of 31 was provided alone, increasing to 54.6% when paired with the cutoff threshold of 30, p=0.0002. This decreased to 33.0% when the FDR of 95% was provided, p=0.003. Likewise, when the abnormality score of 50 was presented alone, desire for MAFI was 31.6%, which increased to 51.3% when paired with the cutoff threshold, which decreased to 36.1% when the FDR was provided, p=0.03.

#### Trust

As seen in Figure 4A and Supplemental Tables 3 and 6, trust in the radiologist and AI after a BI-RADS 1 determination varied depending on whether AI flagged a case and what corresponding information was provided, p=0.0002. Trust in both the radiologist and AI generally decreased when abnormality scores increased. When the FDR of 95% was provided, trust in the radiologist was elevated while trust in AI dropped greatly. Trust in the radiologist was always higher than AI, regardless of experimental condition, p=0.0002.

**Figure 4 A-B.**
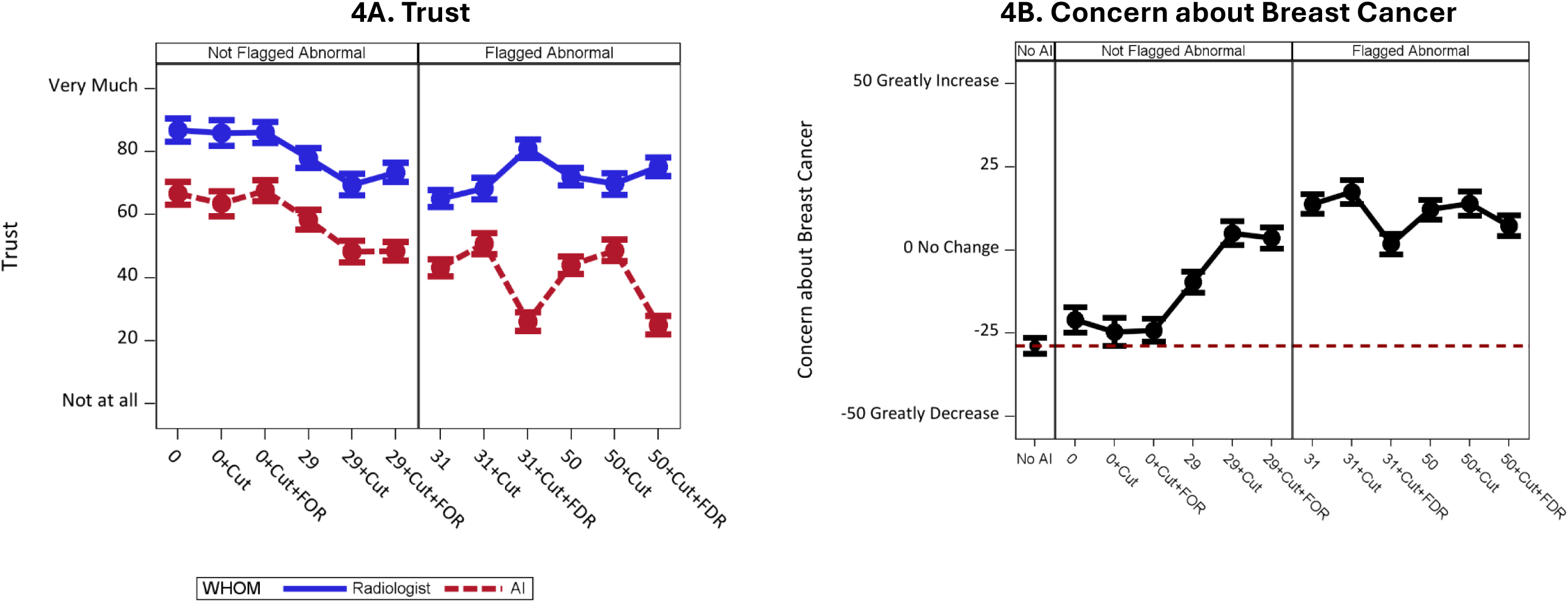
(A) Trust in Radiologist and AI (B) Concern about Breast Cancer. X-Axis is experimental condition. X-Axis is experimental condition, broken down by column panels (No AI (*4B only*), Not Flagged for Abnormality, Flagged for Abnormality). The 12 AI conditions are comprised of four abnormality scores (0, 29, 31, 50) crossed with: 1) the abnormality cutoff threshold of “30” (Cut), and 2) the error rates (FOR, false omission rate, .2%; FDR, false discovery rate, 95%). Y-axis is a magnitude scale of (A) 0 (Not at all) to 100 (Very Much) and (B) a Likert scale (−50 greatly decrease, 0 no change, 50 greatly increase). (A) Blue dots are means with 95% confidence intervals denoting trust in radiologist, and red dots are means with 95% confidence intervals denoting trust in AI. (B) Black dots are means with 95% confidence intervals. Red dashed line is control reference.

#### Concern about breast cancer

As seen in Figure 4B and Supplemental Tables 3 and 6, concern about breast cancer after a BI-RADS 1 determination increased when AI results were included relative to No AI, p=0.0002. Concern about breast cancer increased from −20.9 with No AI (control) to −9.6 when the abnormality score of 29 was presented alone, which increased to 5.1 when paired with the cutoff threshold of 30, which remained at 3.5 when the FOR of 0.2% was included—note, the latter three were relatively high even though AI agreed with the radiologist. Once AI flagged the case as abnormal, concern about breast cancer increased to 13.8 when an abnormality score of 31 was provided alone, increasing to 17.4 when paired with the cutoff threshold of 30, p=0.0002. This decreased to 1.8 when the FDR of 95% was provided, p=0.0002. Likewise, when the abnormality score of 50 was presented alone, concern about breast cancer was 12.1, which increased to 14.0 when paired with the cutoff threshold, which decreased to 7.3 when the FDR was provided, p=0.02.

## DISCUSSION

In this experiment, we demonstrate participants’ hypothetical decision-making behaviors when presented with a BI-RADS 1 radiologist report alongside an AI report vary based on the AI abnormality score, flagging status, and contextual information for interpreting AI triage. Specifically, participants’ consideration of a lawsuit increased with higher AI abnormality scores, and this was mitigated when the FDR or FOR was provided. Furthermore, participants’ requests for additional imaging and follow-up— from the same radiologist, a different radiologist, and the ordering physician—increased as the AI abnormality score increased, though this was often mitigated by providing the FDR (and sometimes FOR).

These findings are important to consider in the context of prior studies probing patient attitudes toward AI in breast imaging. For example, semi-structured interviews by Johansson et al. showed that trust in AI relied upon clear communication about its application and assurance that a radiologist was involved in the assessment process; participants further expressed they would rather tolerate anxiety from frequent callbacks than risk a missed cancer diagnosis.^5^ Mammography alone can already be a source of anxiety, especially when findings are abnormal.^11^ With the introduction of AI results in patient portals, the discordance between a flagged AI report and a BI-RADS 1 radiologist report could be a source of uncertainty and concern for patients that did not exist without AI. Indeed, a recent patient survey found that 91.5% were concerned about AI misdiagnosis.^12^ Our study demonstrates that contextualizing findings by providing the FDR (and sometimes FOR) can reduce the inclination to seek follow-up care or pursue a lawsuit. Providing contextual information to interpret AI findings can be one way to reduce uncertainty and concern.

Additionally, a survey of potential patients by Pesapane et al. found that most approved of AI as a supplementary tool but held both the software developer and radiologist accountable for AI errors.^13^ The “regulatory vacuum” concerning AI in breast imaging—and healthcare more broadly—underscores the need for more robust legal infrastructure to address scenarios when AI errs and, perhaps more importantly, when a radiologist using it errs.^14^ Emerging guidelines have emphasized the importance of appropriate disclosure: Good Machine Learning Practice (GMLP) principles identified by the FDA and collaborating agencies encourage that “users are provided ready access to clear, contextually relevant information that is appropriate for the intended audience.”^15^

Prior research suggests that human factors of AI implementation can impact patient outcomes and legal liability. One study demonstrated that incorrect AI results could worsen radiologist performance; critically, however, implementation strategies mitigated those effects.^16^ A similar consideration translates to the interaction between patient and AI, recognizing that how AI information is presented in patient portals holds downstream implications for imaging volume, healthcare expenditure, and—as shown in the current study—legal liability. Indeed, another vignette experiment demonstrated that when a radiologist provided a false negative report despite AI correctly observing a pathology, mock jurors were more likely to side with the radiologist-defendant if AI accuracy data was (versus was not) provided.^17^ Implementation strategies addressing human factors could reduce existing concerns surrounding trust, transparency, accountability, and legal liability.^4–6^

### Limitations

This study assessed participants’ decision-making behaviors based on hypothetical imaging results bearing no actual consequence on or relationship to the participant, which limits ecological validity. Real-world constraints such as time, financial barriers, and other logistical considerations contribute to patients’ ability to seek second opinions and further imaging. Additionally, the experimental conditions only included a BI-RADS 1 radiologist report with two subthreshold and two supra-threshold AI abnormality scores. Other combinations of concordance and discordance between radiologist and AI were not assessed.

## CONCLUSION

Our study demonstrates that disclosing AI mammography results will impact patients’ interest in pursuing a medical malpractice claim against a radiologist. However, this can be mitigated by providing greater contextual information about an AI system. Moreover, access to AI results impacts patients’ behaviors pertaining to follow-up and checking. AI-enhanced workflows in breast imaging hold significant promise for improving diagnostic accuracy and efficiency, although best practices are needed to inform patients about AI tools used for their care while minimizing the unintended negative consequences.

## Data Availability

All data produced in the present study are available upon reasonable request to the authors

