## Appendix for "Accessing AI mammography reports impacts patient interest in pursuing a medical malpractice claim: The unintended consequences of including AI in patient portals"

Appendices (Supplemental Materials)

### I. Supplemental Tables

**Supplemental Table 1. Hypotheses and Rationale**

| | Hypothesis | Rationale |
|---|---|---|
| 1 | AI-read cases will elevate:<br>• inclination to sue relative to non-AI-read cases if breast cancer is discovered a year later.<br>• checking behavior (second opinion, follow up with same radiologist, follow up with primary care provider, more accurate imaging not otherwise indicated)<br>• concern for having breast cancer | Presence of any AI information will **increase uncertainty** about a radiologist's negative mammography report relative to no-AI information, even for non-flagged cases, especially when more details are provided. |
| 2 | Among the AI-read cases, flagged cases will elevate:<br>• inclination to sue relative to non-AI-read cases if breast cancer is discovered a year later.<br>• checking behavior (second opinion, follow up with same radiologist, follow up with primary care provider, more accurate imaging not otherwise indicated)<br>• concern for having breast cancer | Presence of an AI-flagged determination will **increase uncertainty** about a radiologist's negative mammography report relative to an AI-non-flagged determination. |
| 3 | Among flagged cases, providing the false discovery rate will reduce:<br>• inclination to sue relative to non-AI-read cases if breast cancer is discovered a year later.<br>• checking behavior (second opinion, follow up with same radiologist, follow up with primary care provider, more accurate imaging not otherwise indicated)<br>• concern for having breast cancer | Presence of an AI-flagged determination in the context of a high (95%) false discovery rate (FDR) of said AI system will **decrease uncertainty** about a radiologist's negative mammography report relative to an AI-flagged determination without the FDR. |
| X | Trust in AI | Exploratory |

**Supplemental Table 2. Experimental Conditions**

| Group | BI-RADS 1 (negative finding) | Abnormality score | Flagged Language | Cut-off value of 30 | FOR (0.2%) ✓ FDR (95%) X | Observed N |
|---|---|---|---|---|---|---|
| 1 (Control) | ✓ | | | | | 204 |
| 2 | ✓ | 0 | Not flagged for abnormality | | | 96 |
| 3 | ✓ | 0 | Not flagged for abnormality | ✓ | | 80 |
| 4 | ✓ | 0 | Not flagged for abnormality | ✓ | ✓ | 110 |
| 5 | ✓ | 29 | Not flagged for abnormality | | | 125 |
| 6 | ✓ | 29 | Not flagged for abnormality | ✓ | | 103 |
| 7 | ✓ | 29 | Not flagged for abnormality | ✓ | ✓ | 128 |
| 8 | ✓ | 31 | Flagged for abnormality | | | 151 |
| 9 | ✓ | 31 | Flagged for abnormality | ✓ | | 107 |
| 10 | ✓ | 31 | Flagged for abnormality | ✓ | X | 133 |
| 11 | ✓ | 50 | Flagged for abnormality | | | 149 |
| 12 | ✓ | 50 | Flagged for abnormality | ✓ | | 104 |
| 13 | ✓ | 50 | Flagged for abnormality | ✓ | X | 133 |

**Supplemental Table 3. Comparisons**

| Question | Comparison | Delta | SE | RAW | FDR |
|---|---|---|---|---|---|
| Legal | No AI vs. AI | -0.1918 | 0.05601 | 0.0003 | 0.000557 |
| | No Flag vs. Flag | 0.03618 | 0.112 | 0.3734 | 0.3734 |
| | 31 Cut. Vs. 31 Cut. + FDR | 1.0174 | 0.2697 | <.0001 | 0.00026 |
| | 50 Cut. Vs. 50 Cut. + FDR | 0.616 | 0.2661 | 0.0104 | 0.02 |
| Second Opinion | No AI vs. AI | -0.1681 | 0.06795 | 0.0067 | 0.010247 |
| | No Flag vs. Flag | 1.2289 | 0.1359 | <.0001 | 0.00026 |
| | 31 Cut. Vs. 31 Cut. + FDR | 1.0076 | 0.3057 | 0.0005 | 0.000867 |
| | 50 Cut. Vs. 50 Cut. + FDR | 0.6048 | 0.2857 | 0.0172 | 0.020327 |
| Second Look | No AI vs. AI | -0.761 | 0.076 | <.0001 | 0.000236 |
| | No Flag vs. Flag | 1.1162 | 0.152 | <.0001 | 0.000236 |
| | 31 Cut. Vs. 31 Cut. + FDR | 1.3228 | 0.336 | <.0001 | 0.000236 |
| | 50 Cut. Vs. 50 Cut. + FDR | 0.2756 | 0.3184 | 0.1934 | 0.209517 |
| PCP | No AI vs. AI | -0.1711 | 0.07422 | 0.0107 | 0.02 |
| | No Flag vs. Flag | 0.9082 | 0.1484 | <.0001 | 0.000236 |
| | 31 Cut. Vs. 31 Cut. + FDR | 0.798 | 0.3581 | 0.013 | 0.016095 |
| | 50 Cut. Vs. 50 Cut. + FDR | 0.174 | 0.3577 | 0.3134 | 0.325936 |
| Follow-up Imaging | No AI vs. AI | -1.2918 | 0.08406 | <.0001 | 0.000217 |
| | No Flag vs. Flag | 1.0921 | 0.1681 | <.0001 | 0.000217 |
| | 31 Cut. Vs. 31 Cut. + FDR | 0.8893 | 0.3064 | 0.0019 | 0.003088 |
| | 50 Cut. Vs. 50 Cut. + FDR | 0.6244 | 0.312 | 0.0228 | 0.025774 |
| Trust | Interaction Effect | | | <.0001 | 0.000217 |
| | Main Effect (Rad vs. AI) | | | <.0001 | 0.0002 |
| Concern | No AI vs. AI | 51.4038 | 0.6098 | <.0001 | 0.0002 |
| | No Flag vs. Flag | 26.3817 | 1.2197 | <.0001 | 0.0002 |
| | 31 Cut. Vs. 31 Cut. + FDR | 15.5666 | 2.8734 | <.0001 | 0.0002 |
| | 50 Cut. Vs. 50 Cut. + FDR | 6.6244 | 2.8973 | 0.0112 | 0.02 |

**Supplemental Table 4. Participants Considering Lawsuit by Condition**

| Condition | *M (%)* | *[95% CI]* |
|---|---|---|
| 0 | 46.9 | [37.1, 56.9] |
| 0+Cut | 43.8 | [33.3, 54.8] |
| 0+Cut+FOR | 40.0 | [31.3, 49.4] |
| 29 | 44.8 | [36.3, 53.6] |
| 29+Cut | 45.6 | [36.3, 55.3] |
| 29+Cut+FOR | 47.7 | [39.2, 56.3] |
| 31 | 60.7 | [52.6, 68.2] |
| 31+Cut | 56.1 | [46.6, 65.2] |
| 31+Cut+FDR | 31.6 | [24.3, 40.0] |
| 50 | 48.3 | [40.4, 56.3] |
| 50+Cut | 51.9 | [42.4, 61.4] |
| 50+Cut+FDR | 36.8 | [29.1, 45.4] |
| No AI | 39.7 | [33.2, 46.6] |

**Supplemental Table 5. Participants Requesting Second Opinion, Second Look, Visit with Primary Care Physician, Follow-Up Imaging by Condition**

| | Second Opinion | | Second Look | | Visit with PCP | | Follow-Up Imaging | |
|---|---|---|---|---|---|---|---|---|
| Condition | *M (%)* | *[95% CI]* | *M (%)* | *[95% CI]* | *M (%)* | *[95% CI]* | *M (%)* | *[95% CI]* |
| 0 | 11.5 | [6.5,19.5] | 10.6 | [5.8, 18.7] | 14.3 | [8.5, 23.1] | 7.4 | [3.6, 14.7] |
| 0+Cut | 20.0 | [12.6, 30.2] | 14.3 | [8.1, 24.0] | 20.6 | [12.8, 31.3] | 7.8 | [3.5, 16.3] |
| 0+Cut+FOR | 17.6 | [11.5, 26.0] | 10.5 | [5.9, 17.9] | 23.8 | [16.5, 33.0] | 6.6 | [3.2, 13.2] |
| 29 | 27.2 | [20.1, 35.7] | 21.8 | [15.1, 30.5] | 37.6 | [28.7, 47.4] | 13.3 | [8.2, 20.9] |
| 29+Cut | 68.0 | [58.4, 76.3] | 45.1 | [33.9, 56.7] | 67.7 | [55.2, 78.2] | 25.3 | [16.8, 36.4] |
| 29+Cut+FOR | 62.5 | [53.8, 70.5] | 40.7 | [31.1, 51.0] | 58.0 | [47.4, 67.8] | 39.6 | [30.6, 49.4] |
| 31 | 70.2 | [62.4, 77.0] | 61.5 | [51.9, 70.4] | 64.9 | [53.4, 74.9] | 47.5 | [38.6, 56.5] |
| 31+Cut | 81.3 | [72.8, 87.6] | 70.6 | [58.7, 80.2] | 73.3 | [60.8, 83.0] | 54.6 | [43.4, 65.3] |
| 31+Cut+FDR | 61.4 | [52.8, 69.3] | 39.0 | [30.0, 48.9] | 55.3 | [45.2, 65.0] | 33.0 | [24.9, 42.4] |
| 50 | 68.9 | [61.0, 75.9] | 44.6 | [34.8, 54.8] | 59.2 | [47.9, 69.7] | 31.6 | [23.9, 40.6] |
| 50+Cut | 74.0 | [64.8, 81.6] | 57.4 | [45.4, 68.5] | 63.0 | [49.4, 74.7] | 51.3 | [40.2, 62.3] |
| 50+Cut+FDR | 60.9 | [52.4, 68.8] | 50.5 | [40.7, 60.3] | 58.8 | [48.1, 68.8] | 36.1 | [27.2, 46.1] |
| No AI | 8.3 | [5.2, 13.0] | 9.8 | [6.4, 14.9] | 19.8 | [14.7, 26.0] | 7.2 | [4.3, 11.8] |

**Supplemental Table 6. Participants' Concern About Breast Cancer, Trust in AI, Trust in Radiologist by Condition**

| | Concern About Breast Cancer | | Trust in AI | | Trust in Radiologist | |
|---|---|---|---|---|---|---|
| Condition | *M (%)* | *[95% CI]* | *M (%)* | *[95% CI]* | *M (%)* | *[95% CI]* |
| 0 | 29.1 | [24.6, 33.6] | 66.8 | [62.4, 71.1] | 86.9 | [82.5, 91.3] |
| 0+Cut | 25.4 | [20.5, 30.3] | 63.5 | [58.7, 68.3] | 86.0 | [81.2, 90.7] |
| 0+Cut+FOR | 25.9 | [21.8, 30.0] | 67.7 | [63.7, 71.8] | 86.1 | [82.0, 90.1] |
| 29 | 40.4 | [36.6, 44.2] | 58.4 | [54.5, 62.3] | 78.0 | [74.2, 81.8] |
| 29+Cut | 55.1 | [50.8, 59.3] | 48.3 | [44.1, 52.5] | 69.6 | [65.4, 73.7] |
| 29+Cut+FOR | 53.5 | [49.7, 57.4] | 48.5 | [44.7, 52.2] | 73.4 | [69.7, 77.2] |
| 31 | 63.8 | [60.3, 67.4] | 43.2 | [39.7, 46.6] | 65.1 | [61.7, 68.6] |
| 31+Cut | 67.4 | [63.2, 71.6] | 50.8 | [46.7, 54.9] | 68.3 | [64.2, 72.4] |
| 31+Cut+FDR | 51.8 | [48.1, 55.6] | 26.1 | [22.4, 29.7] | 80.9 | [77.2, 84.6] |
| 50 | 62.1 | [58.6, 65.7] | 44.0 | [40.5, 47.5] | 72.1 | [68.6, 75.6] |
| 50+Cut | 64.0 | [59.7, 68.2] | 48.7 | [44.5, 52.9] | 69.9 | [65.7, 74.0] |
| 50+Cut+FDR | 57.3 | [53.6, 61.1] | 25.0 | [21.3, 28.7] | 75.2 | [71.5, 78.9] |
| No AI | 21.3 | [18.3, 24.3] | | | | |

### II. Vignettes

**All Conditions (1-13)**

Imagine you just returned from your annual screening mammogram and you receive an email notification that the report of your mammogram results is available in your online patient portal. The report is from the radiologist. A radiologist is a doctor who specializes in reading medical images, such as mammograms.

You log in and read the following:

**Radiologist Final Report**

**FINDINGS:**
There are no suspicious masses, suspicious calcifications, or secondary signs of malignancy within either breast.

**IMPRESSION:**
No mammographic evidence of malignancy.

**BIRADS 1**: Negative

**Follow-up Recommendation**: Annual Screening

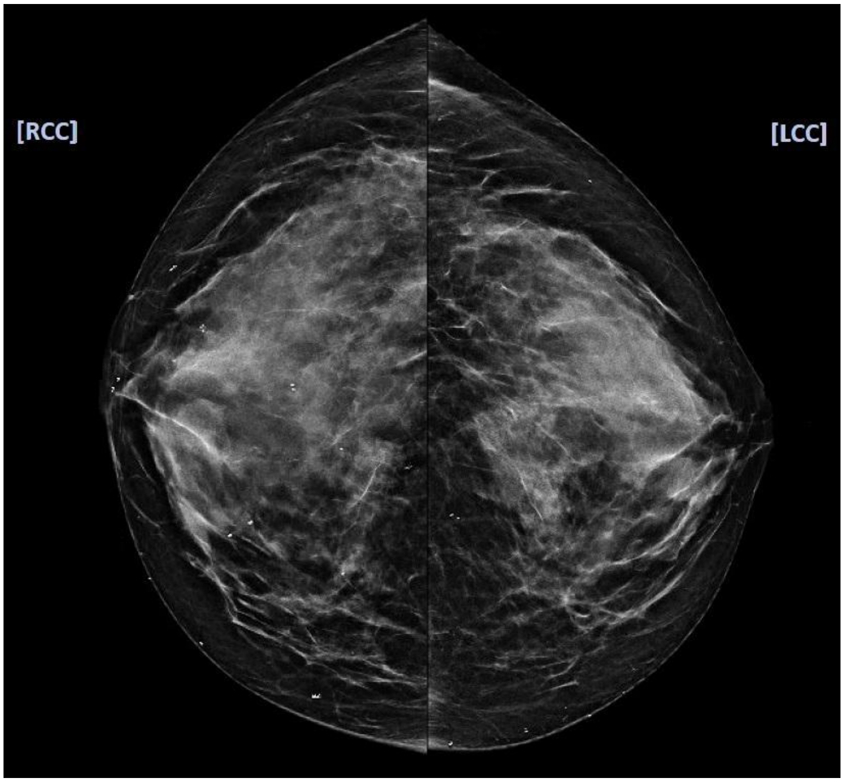

**AI Conditions (2-13). Note, for brevity, 29+Cut+FOR is used as an example.**

### FDA-Cleared AI Tool Report

**Patient's Abnormality Score**: 29 (Not flagged for abnormality)

Note: Possible abnormality scores range from 0 to 100. The threshold score for the AI tool flagging possible cancer is 30 or higher.

Note: The False Omission Rate for this AI tool is 0.2%. In other words, for every 1000 cases that AI finds no evidence of abnormality, it misses 2 cases that are actually cancer (and the other 998 are negative for cancer).

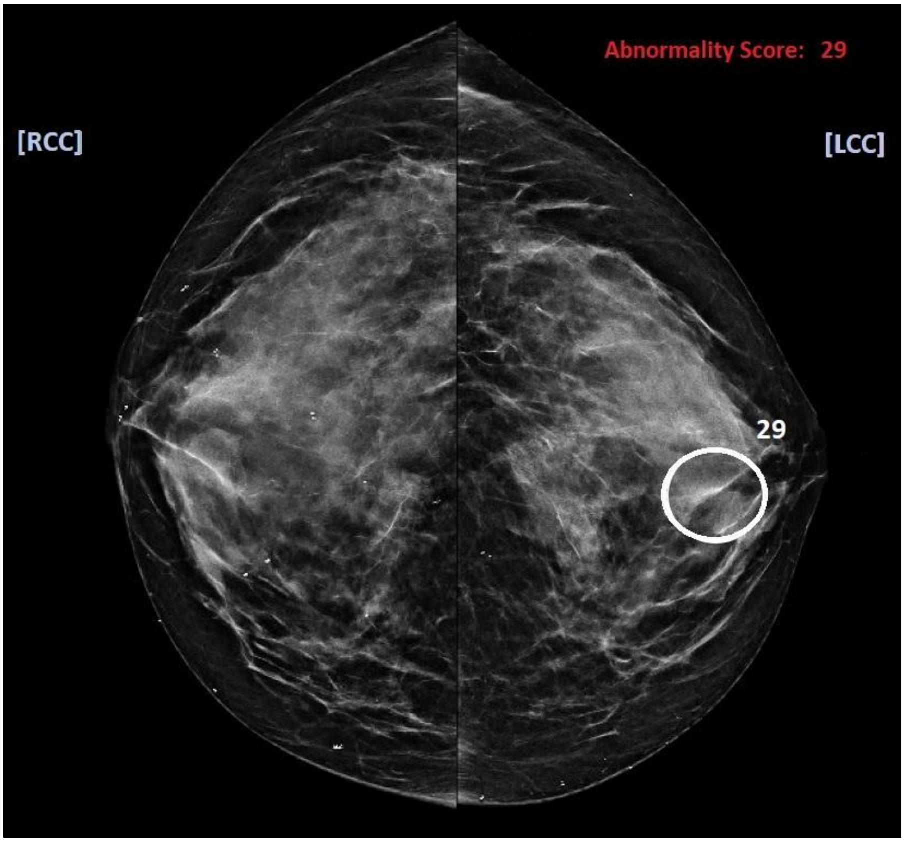
